# The Association Between the Rate of Angiotensin-Converting Enzyme Inhibitors and Angiotensin Receptor Blockers Use and the Number of Covid-19 Confirmed Cases and Deaths in the United States: Geospatial Study

**DOI:** 10.1101/2020.05.31.20118802

**Authors:** Kyle Johnson, Maedeh Khayyat-Kholghi, Blake Johnson, Larisa G. Tereshchenko

**Affiliations:** Knight Cardiovascular Institute, Oregon Health & Science University, Portland, OR; Flexport, San Francisco, CA

**Keywords:** ACEI, ARB, COVID-19, coronavirus

## Abstract

**Background:** The novel coronavirus SARS-Cov2 uses the angiotensin-converting enzyme 2 (ACE2) receptor as an entry point to the cell. Cardiovascular disease (CVD) is a risk factor for the novel coronavirus disease (Covid-19) with poor outcomes. We hypothesized that the rate of ACE inhibitor (ACEI) and angiotensin receptor blocker (ARB) use is associated with the rate of Covid-19 confirmed cases and deaths.

**Methods:** We conducted a geospatial study using publicly available county-level data. The Medicare ACEI and ARB prescription rate was exposure. The Covid-19 confirmed case and death rates were outcomes. Spatial autoregression models were adjusted for the percentage of Black residents, children, residents with at least some college degree, median household income, air quality index, CVD hospitalization rate in Medicare beneficiaries, and CVD death rate in a total county population.

**Findings:** After adjustment for confounders, the ACEI use rate did not associate with Covid-19 confirmed case rate (direct county-own effect +0.11 %; 95%CI -0.31 to 0.53; P=0.600, and indirect spillover effect -0.53 %; 95%CI -3.89 to 2.84; P=0.760). The ARB use rate was associated with increased Covid-19 confirmed case rate (direct county-owned effect +0.12 %; 95%CI 0.05-0.19; P=0.002, and indirect spillover effect -0.33 %; 95%CI -2.11 to 1.44; P=0.714). Sensitivity analysis indicated an absence of significant reverse causality bias for analyses with Covid-19 confirmed case rate, but not death rate outcome.

**Interpretation:** Our results highlight the safety of ACEI use for patients with clinical indications for ACEI use. However, an increase in ARB use by 1% was associated with a 0.12 % increase in Covid-19 confirmed cases. The use of ARB, due to known ACE2 upregulation, may facilitate SARS-CoV-2 entry into target cells and increase infectivity. Cluster-randomized controlled trial is warranted to answer the question of whether the replacement of ARB by ACEI may reduce the Covid-19 confirmed case rate.

**Funding:** LGT was supported in part by the National Institute of Health (HL118277).

## Introduction

The novel coronavirus disease 2019 (Covid-19) caused by SARS-Cov2 virus was named a pandemic officially by the World Health Organization on March 11^th^, 2020.^1^ It has been shown that SARS-Cov2 uses the angiotensin-converting enzyme 2 (ACE2) receptor as an entry point into a cell.^2^

With the ACE2 receptor acting as a binding site for SARS-Cov2, the Renin-Angiotensin-Aldosterone System (RAAS), and the medications affecting it become important points of discussion.^3^ Angiotensin-converting enzyme inhibitors (ACEI) and angiotensin receptor blockers (ARB) are two classes of medications widely used in patients with hypertension, diabetes, cardiovascular disease (CVD), and latent or manifest left ventricular dysfunction. Hypertension, diabetes, and CVD emerged as risk factors for severe Covid-19 cases and deaths.^4^ Previous experiments showed that ACE2 expression is associated with susceptibility to SARS-Cov infection.^5^

Notably, available clinical studies consider ACEI and ARB together. However, the effects of ACEI and ARB on ACE2 levels and activity differ.^3^ While there is strong evidence that ARBs increase ACE2 expression^6-11^ and augment ACE2 activity^6,12^, only Ferrario et al^13^ showed that ACEIs increase ACE2 expression, whereas other studies showed no effect of ACEI on ACE2 activity.^14-16^

The consensus is reached by all international cardiac societies about the importance of the continuation of ACEI and ARB use in Covid-19 pandemic. However, it remains unknown whether clinically indicated use of ACEI and ARB improve or worsen infectivity or the course of Covid-19. In this rapidly growing pandemic, time is of an essence. To address an urgent need, we conducted a geospatial study. We hypothesized that in the geospatial analysis, the rate of ACEI and ARB use are associated with the number of confirmed Covid-19 cases and deaths in the United States (US).

## Methods

We conducted a geospatial disease mapping study using publicly available county-level data. The study was reviewed by the Oregon Health & Science University Institutional Review Board and assigned a determination of Not Human Research. We provided the study dataset and STATA (StataCorp, College Station, Texas) code at https://github.com/Tereshchenkolab/geospatial, allowing future replication and update of the study results as COVID-19 pandemic is unfolding.

### Geographical framework

An individual county in the US was an observation unit in this study. We used a Federal Information Processing Standard (FIPS) county code to link the data. Data with missing FIPS codes were excluded from the study. Geographic information about each county was obtained from the cartographic boundary files (shapefiles) provided by the US Census Bureau’s MAF/TIGER geographic database.^17^

### Exposure: rate of ACEI and ARB use by the Medicare Part D beneficiaries

We used the 2017 Centers for Medicare & Medicaid Services (CMS) public dataset, the Medicare Provider Utilization and Payment Data: Part D Prescriber Public Use File, with information on prescription drugs prescribed by individual physicians and other health care providers and paid for under the Medicare Part D Prescription Drug Program in 2017.^18^ The dataset included the total number of prescriptions that were dispensed (total day supply), which include original prescriptions and any refills, and, therefore, reflects ACEI and ARB usage.

The Medicare dataset only includes city and state information and not county information. In order to map the Medicare prescription data to their corresponding FIPS Code, we used the Google Geocoding API.^19^ We then loaded the Medicare data and geocoded data into the SQLite database to produce the final datasets with prescription counts per county. Prescriptions from county-equivalents (independent cities) were manually matched with their corresponding FIPS code. Medicare prescriptions with misspelled cities or prescriptions that lacked valid city and state descriptions, if unable to be assigned, were excluded. Excluded prescriptions accounted for <0.01% of the data.

We calculated a drug class use rate as a sum of total day supply in a county for all drugs comprising a particular class (Table 1), normalized by the total county population estimate. We used the US Census Annual Resident Population Estimates for July 1^st^, 2019.

**Table 1.**
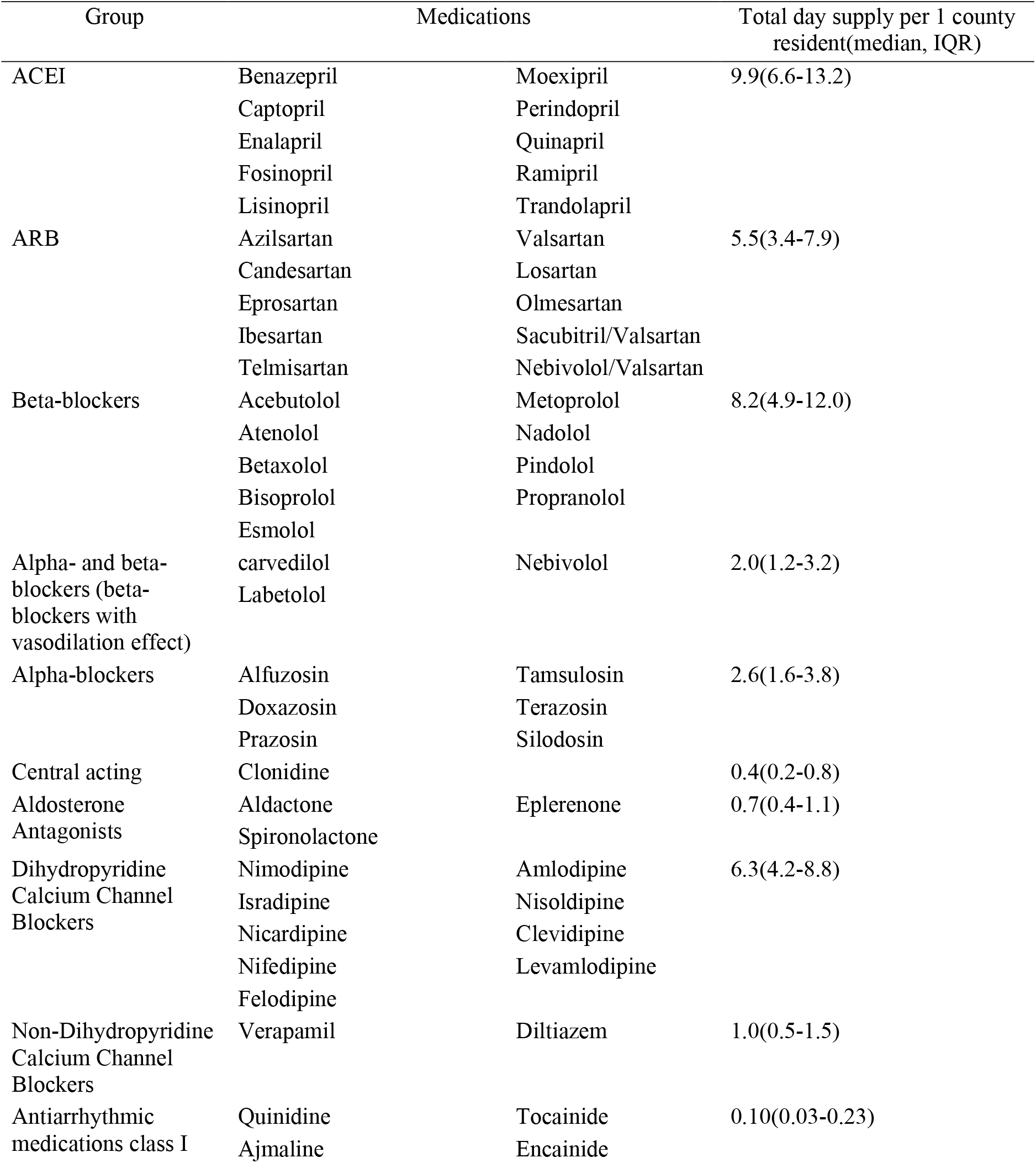
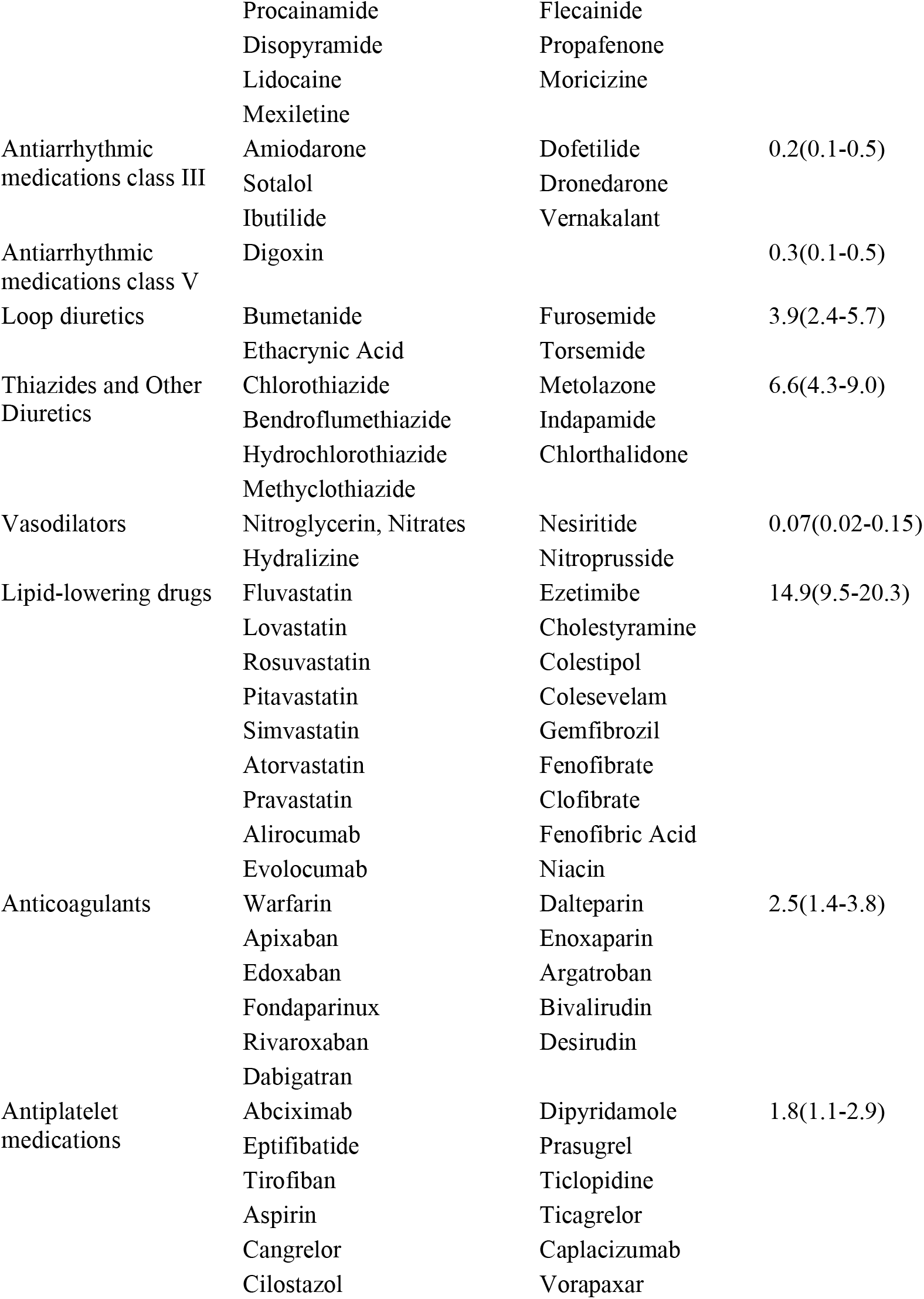
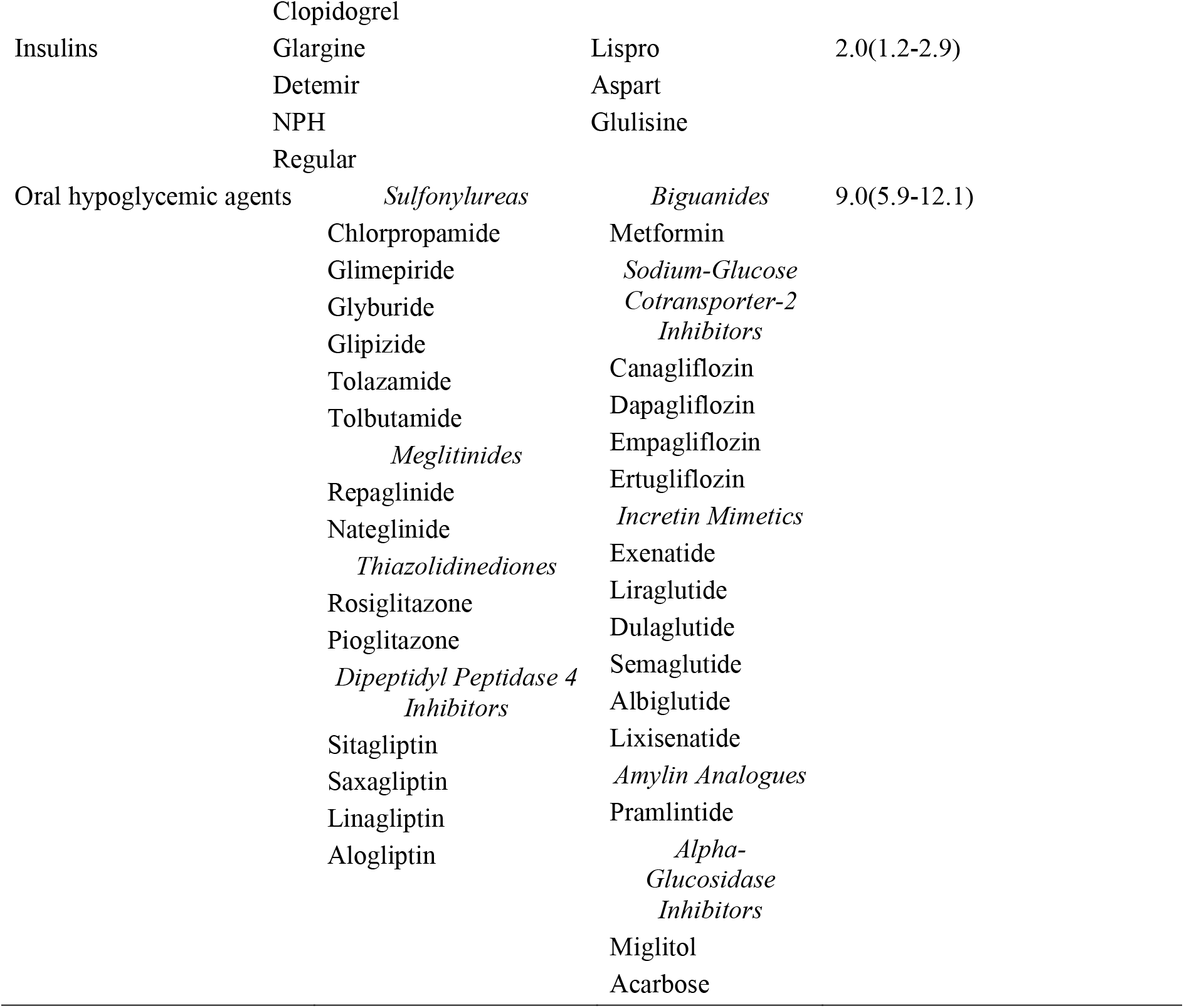
List of studied medications and average yearly total day supply per 1 county resident

### Outcomes: COVID-19 confirmed cases and deaths

The primary outcome was confirmed Covid-19 cases. The secondary outcome was Covid-19 deaths. We imported the raw COVID-19 data from the Johns Hopkins GitHub repository.^20^ The number of confirmed COVID-19 cases and deaths in each county as reported for June 11^th^, 2020 was divided by the total population in each county (2019 county population estimate) and multiplied by 100,000 to convert to cases and deaths per 100,000 population.

### Covariates: Population characteristics

#### Demographic and socioeconomic characteristics

We used the US Census County Population Estimates, released in March 2020, and included reported deaths and births in period July 1^st^, 2018 to June 30^th^, 2019.^21^ Due to known negative impact of Covid-19 on the population of nursing homes and prisons/jails, we included July 1^st^, 2019 Group Quarters total population estimate. Group Quarters Facilities include correctional facilities for adults, nursing homes, college/university student housing, military quarters, and group homes. Group Quarters data was gathered from an estimated 20,000 randomly selected facilities. Data was then collected through resident interviews of these selected facilities using the American Community Survey conducted by the US Census Bureau.^21^ The total 2019 county population estimate^21^ normalized all demographic characteristics.

To characterize socioeconomic characteristics, we used the 2018 median household income expressed as a percent of the state total, and percent of the total population in poverty, as reported by the Economic Research Service of the US Department of Agriculture.^22^ We also used the data compiled by the County Health Rankings & Roadmaps program, which is a collaboration between the Robert Wood Johnson Foundation and the University of Wisconsin Population Health Institute.^23^

#### Cardiovascular disease prevalence and severity

To characterize CVD prevalence and severity, we used the Centers for Disease Control and Prevention (CDC) estimates^24^ of total CVD death rate per 100,000 population (2016-18), total CVD hospitalizations (2015-17) per 1,000 Medicare beneficiaries, heart failure (HF) death rate per 100,000 population (2016-18), HF hospitalization rate per 1,000 Medicare beneficiaries (2015-17), coronary heart disease (CHD) death rate per 100,000 population (2016-18), CHD hospitalization rate per 1,000 Medicare beneficiaries (2015-17), and age-adjusted diabetes percentage in adults (age > 20y). This data was obtained from the Interactive Atlas of Heart Disease and Stroke, published by the CDC. Within this atlas, death rates were gathered from the Deaths National Vital Statistics Program, hospitalization rates were gathered from the Centers for Medicare and Medicaid Services Medicare Provider Analysis and Review file Part A, and diabetes percentages were collected from the Division of Diabetes Translation.^24^

To characterize the use of cardiovascular medications, we calculated the rate of cardiovascular medications use, which included original prescriptions and any refills (total day supply), as reported in the 2017 CMS Part D Medicare Prescriber Public Use File.^18^ We considered the total day supply data for 20 medication groups (Table 1): ACEI, ARB, beta-blockers, alpha-and-beta-blockers, alpha-blockers, class I, III, and V antiarrhythmic medications, dihydropyridine, and non-dihydropyridine calcium channel blockers, aldosterone antagonists, central acting antihypertensive medications, vasodilators, diuretics, lipid-lowering drugs, insulins, and oral hypoglycemic agents, anticoagulants and antiplatelet medications. We normalized the cardiovascular medications day supply for each county by the 2019 county population estimate.^21^

### Statistical analyses

Data are summarized as mean ± standard deviation or as the median and interquartile range (IQR) if non-normally distributed. A paired *t*-test was used to compare an average rate of ACEI and ARB use.

As Covid-19 is a contagious disease, confirmed cases and deaths in neighboring counties are spatially correlated. Therefore, we used spatial autoregression model^25^ that allows modeling the spatial dependence among the outcomes, covariates, and among unobserved errors.^26^ The spatial autoregression model used the generalized spatial two-stage (method-of-moment), least-squares estimator.^27^ The model included spatial lags of the outcome variable, spatial lags of covariates, and spatially autoregressive errors. The lag operator was a spatial weighting (inverse-distance) matrix, which summarized spatial relationships between counties, based on the distance between county centroids. Weighting matrix was scaled so that its largest eigenvalue is 1, which guarantees nonsingularity in the model estimation. We constructed cross-sectional spatial autoregressive models. The estimator treated the errors as heteroskedastic, thus relaxing the assumption that errors represent identically distributed disturbance.

We conducted the Moran test to determine whether exposure, outcome, and covariate variables are spatially dependent.

First, we constructed unadjusted spatial autoregression models, to investigate a geospatial association of the county population characteristics with the ACEI and ARB use rate, calculated as follows:

ACEI use rate = ln (1+total ACEI day supply/county population).

ARB use rate = ln (1+total ARB day supply/county population).

Each model included spatial lags of the outcome variable (ACEI or ARB use rate, one-by-one), spatial lags of the test’s predictor variable (demographic, socioeconomic, and CVD prevalence characteristics, one-by-one), and spatially autoregressive errors (reflecting unobserved factors).

Next, we constructed unadjusted and bivariate spatial autoregression models, to evaluate a geospatial association of the county population characteristics with the rate of Covid-19 confirmed cases and deaths. To normalize the distribution of the outcome variables, and to improve the interpretability of models, we transformed outcome variables as follows:

Covid-19 confirmed case rate = ln (1+ confirmed Covid-19 cases/100,000 population); Covid-19 death rate = ln (1+ confirmed Covid-19 deaths/100,000 population).

Each model included spatial lags of the outcome variable (Covid-19 confirmed case rate and death rate, one-by-one), spatial lags of the tested predictor variable (demographic and socioeconomic characteristics, cardiovascular risk factors and CVD prevalence, and the rate of

ACEI or ARB use, one-by-one), and spatially autoregressive errors (unobserved impacts). The bivariate model included both predictors (ACEI and ARB use) together.

Finally, we constructed adjusted spatial autoregression models to answer the main study question: whether there is an independent association of ACEI and ARB use rate with Covid-19 confirmed case and death rate. The selection of covariates for adjustment was guided by confounders observed in this study and model fit. We adjusted for demographic and socioeconomic characteristics of a county population (percentage of Black non-Hispanic county residents, percentage of a county residents younger than 18 years of age, percentage of residents with at least some college degree, median household income as a percent of state total), air quality index, as well as CDC-reported CVD hospitalization rate in Medicare beneficiaries, and CVD death rate in a total county population. Each model included spatial lags of the outcome variable (Covid-19 confirmed cases and deaths, one-by-one), spatial lags of the tested predictor variable (rate of ACEIs or ARBs usage, one-by-one), spatial lags of the selected (listed above) 7 covariates (altogether) and spatially autoregressive errors (unobserved influences).

#### Sensitivity analyses

Cross-sectional geospatial analysis is susceptible to reverse causality bias. It is well-documented that patients with CVD and cardiovascular risk factors (hypertension, diabetes) have a higher rate of Covid-19 confirmed cases and deaths. The rate of ACEIs and ARBs use indirectly indicates CVD prevalence and severity. While we adjusted our models for the broad range of confounders, including CVD mortality in a total county population, and CVD hospitalization rate among Medicare beneficiaries, reverse causality remained of concern. To assess the possibility and extent of reverse causality bias, we constructed the aforementioned spatial autoregression models for the use rate of other cardiovascular medications, for each class of drugs separately, one-by-one.

As it is known that the angiotensin receptor–neprilysin inhibitors are used as second-line therapy for the treatment of advanced HF, we conducted a sensitivity analysis with pure ARB class of medications to minimize the reverse causality bias.

## Results

### Rate of ACEI and ARB use, and their association with population characteristics

We analyzed the data of 3,141 counties and county-equivalents in the 50 states and the District of Columbia. ACEI were the second most ubiquitous medications, surpassed only by lipid-lowering drugs (Table 1). Average county characteristics are reported in Table 2. Figure 1 shows the ACEI and ARB total day supply rates across the US. On average, the total day supply rate was higher for ACEI than for ARB (10.3±6.5 vs. 6.0±4.6 days; *P*<0.0001), as shown in Figure 1C. The Moran test indicated that the rates of ACEI and ARB use were spatially dependent (*P*<0.0001).

**Table 2.**
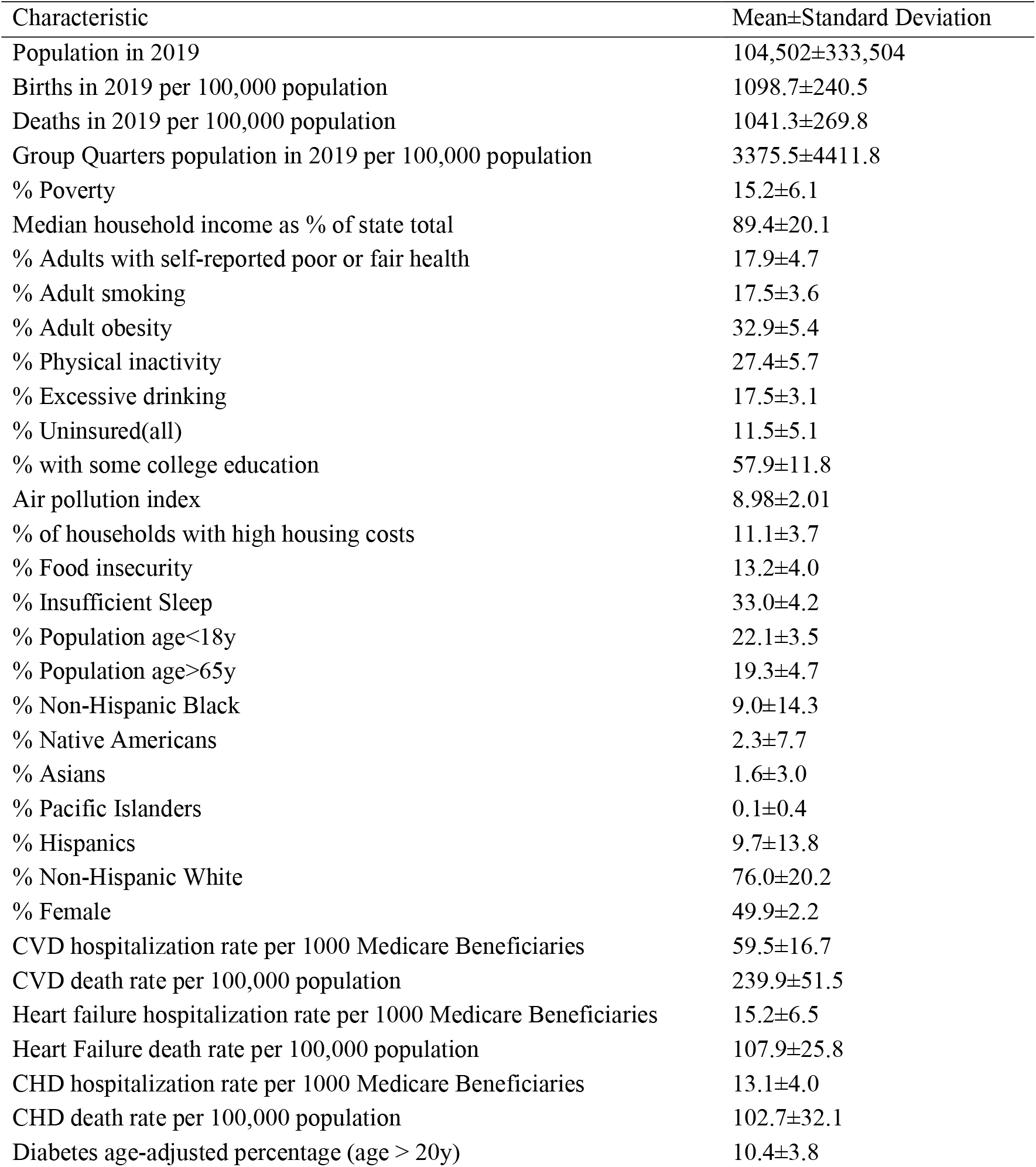
Average characteristics of counties

| Characteristic | Mean±Standard Deviation |
| --- | --- |
| Population in 2019 | 104,502±333,504 |
| Births in 2019 per 100,000 population | 1098.7±240.5 |
| Deaths in 2019 per 100,000 population | 1041.3±269.8 |
| Group Quarters population in 2019 per 100,000 population | 3375.5±4411.8 |
| % Poverty | 15.2±6.1 |
| Median household income as % of state total | 89.4±20.1 |
| % Adults with self-reported poor or fair health | 17.9±4.7 |
| % Adult smoking | 17.5±3.6 |
| % Adult obesity | 32.9±5.4 |
| % Physical inactivity | 27.4±5.7 |
| % Excessive drinking | 17.5±3.1 |
| % Uninsured(all) | 11.5±5.1 |
| % with some college education | 57.9±11.8 |
| Air pollution index | 8.98±2.01 |
| % of households with high housing costs | 11.1±3.7 |
| % Food insecurity | 13.2±4.0 |
| % Insufficient Sleep | 33.0±4.2 |
| % Population age<18y | 22.1±3.5 |
| % Population age>65y | 19.3±4.7 |
| % Non-Hispanic Black | 9.0±14.3 |
| % Native Americans | 2.3±7.7 |
| % Asians | 1.6±3.0 |
| % Pacific Islanders | 0.1±0.4 |
| % Hispanics | 9.7±13.8 |
| % Non-Hispanic White | 76.0±20.2 |
| % Female | 49.9±2.2 |
| CVD hospitalization rate per 1000 Medicare Beneficiaries | 59.5±16.7 |
| CVD death rate per 100,000 population | 239.9±51.5 |
| Heart failure hospitalization rate per 1000 Medicare Beneficiaries | 15.2±6.5 |
| Heart Failure death rate per 100,000 population | 107.9±25.8 |
| CHD hospitalization rate per 1000 Medicare Beneficiaries | 13.1±4.0 |
| CHD death rate per 100,000 population | 102.7±32.1 |
| Diabetes age-adjusted percentage (age > 20y) | 10.4±3.8 |

**Figure 1.**
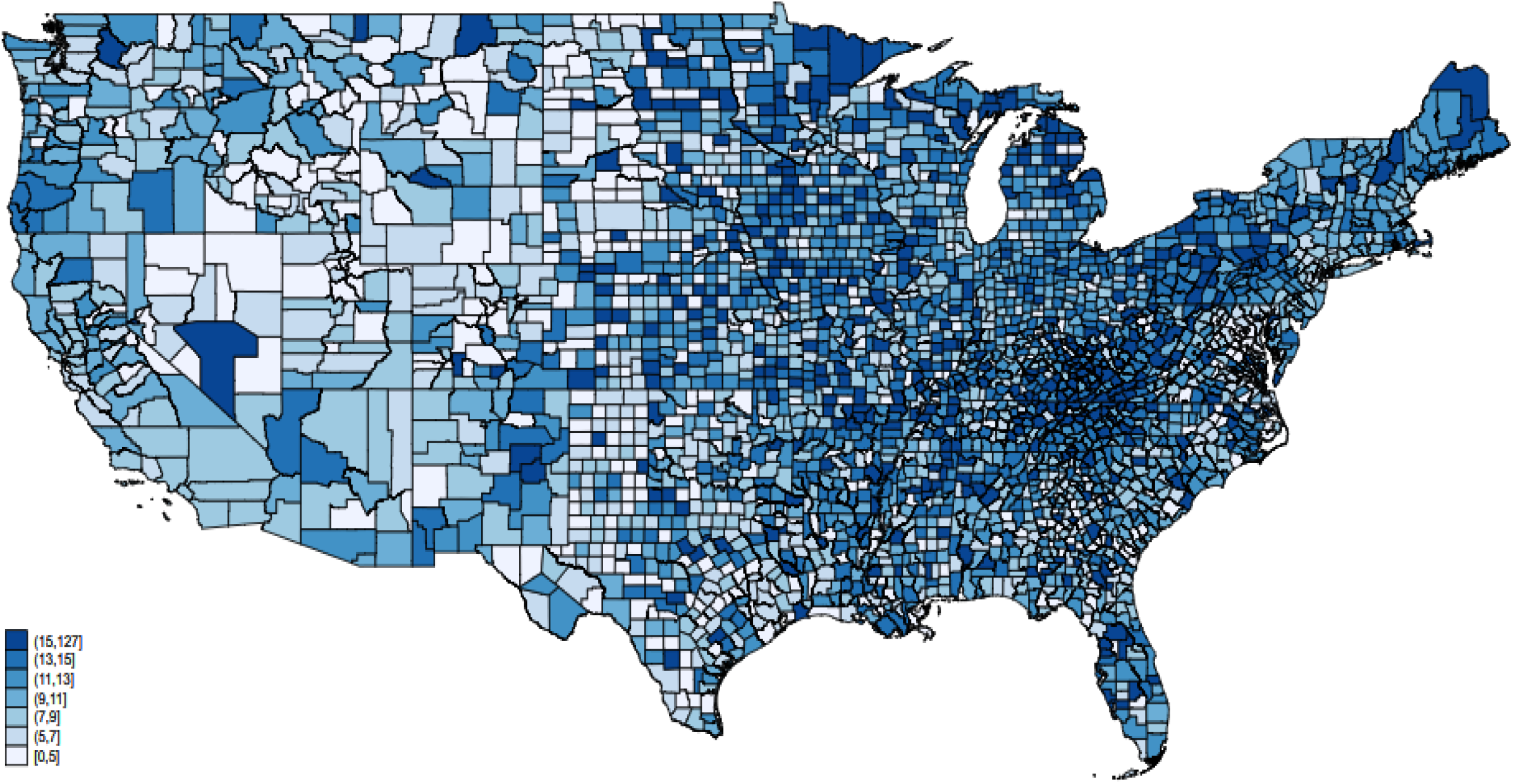
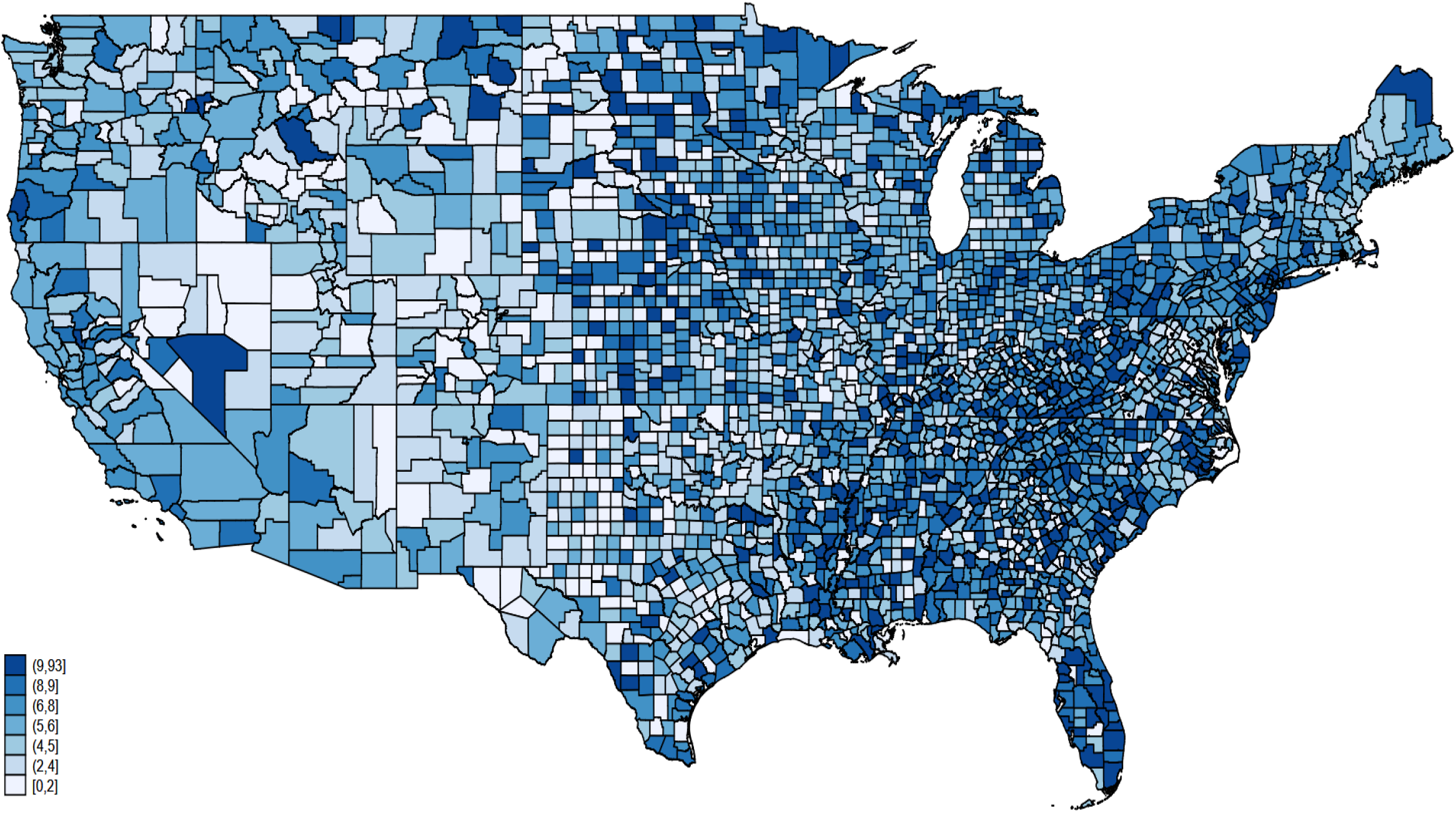
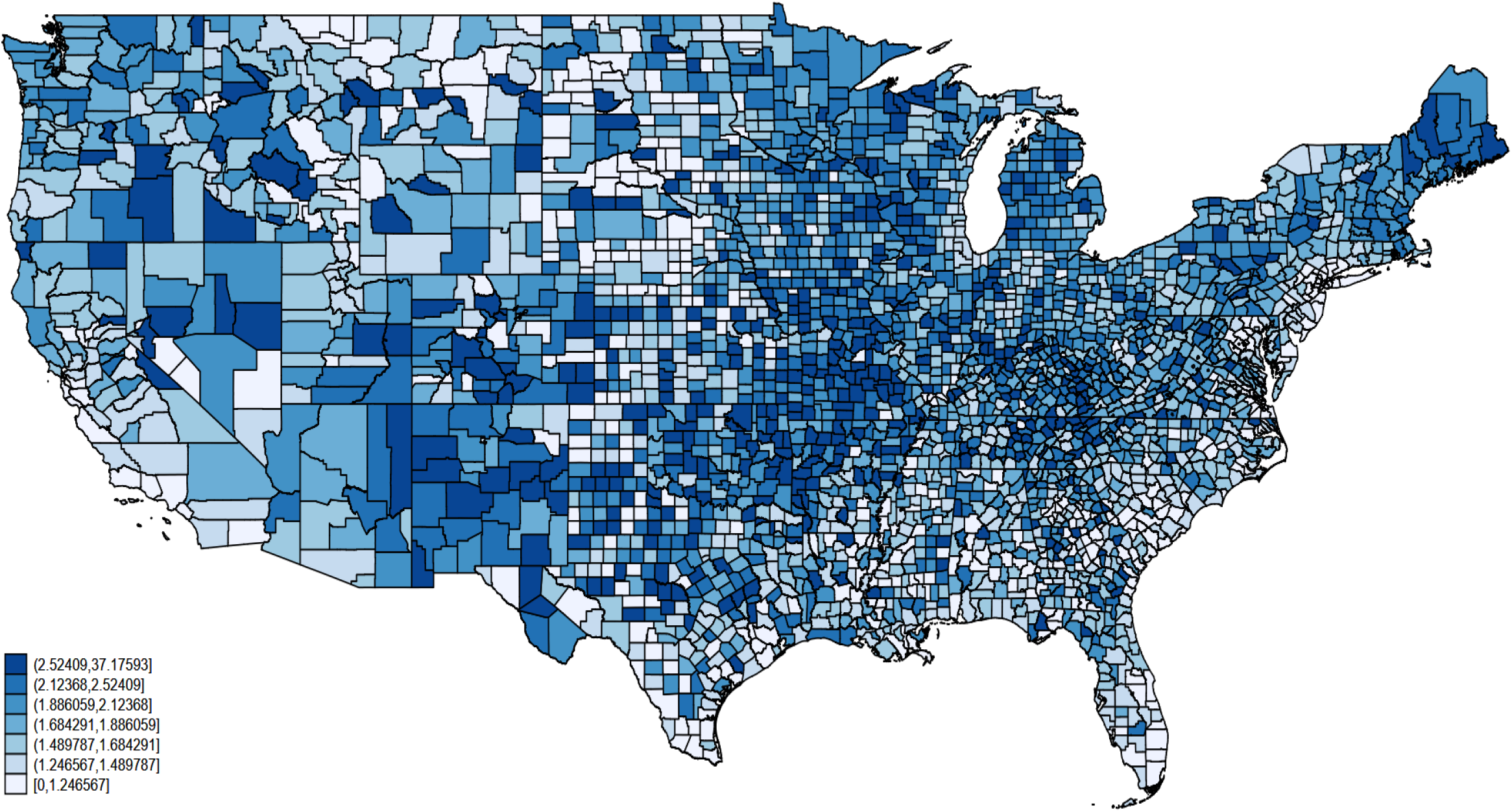
ACEIs (**A**) and ARBs (**B**) total day supply rate and their ratio (**C**).

In unadjusted spatial autoregression analysis (Table 3), as expected, CVD prevalence, general demographic characteristics, uninsured rate, and air quality were associated with the use of both ACEI and ARB. A higher percentage of adults above 65 y of age was associated with higher use of ACEI, but not ARB. A higher percentage of Asians was associated with the use of ARB, but not ACEI.

**Table 3.** Unadjusted impact on geospatial distribution of ACEI and ARB use

| Impact factor | ACEI use rate |  |  |  | ARB use rate |  |  |  |
| --- | --- | --- | --- | --- | --- | --- | --- | --- |
|  | Direct (county-own) effect |  | Indirect (spillover) effect |  | Direct (county-own) effect |  | Indirect (spillover) effect |  |
| Per 1% rate increase | Marginal effect (95%CI) | P-value | Marginal effect (95%CI) | P-value | Marginal effect (95%CI) | P-value | Marginal effect (95%CI) | P-value |
| Births in 2019 | **+0.24(0.07-0.42)** | **0.006** | -1.91(-10.01-6.2) | 0.645 | **+0.33(0.16-0.51)** | **<0.0001** | -2.5(-8.7-3.9) | 0.417 |
| Deaths in 2019 | **+0.41(0.17-0.65)** | **0.001** | **-1.56(-2.85 to -0.26)** | **0.018** | **+0.32(0.14-0.50)** | **<0.0001** | **-1.24(-1.91 to -0.56)** | **<0.0001** |
| GQ population 2019 | -0.003(-0.039-0.033) | 0.867 | **-0.38(-0.50 to -0.26)** | **<0.0001** | +0.014(-0.021-0.050) | 0.426 | -2.75(-13.99-8.49) | 0.632 |
| Poverty | **+0.01(0.004-0.02)** | **0.001** | -0.27(-0.82-0.29) | 0.348 | +0.004(-0.005-0.01) | 0.383 | -0.03(-0.14-0.08) | 0.625 |
| Median HH income | **-0.005(-0.006 to -0.003)** | **<0.0001** | +0.009(-0.01-0.03) | 0.372 | -0.001(-0.003-0.0004) | 0.176 | -0.05(-0.28-0.19) | 0.687 |
| Poor/Fair Health | +1.26(-10.8-13.7) | 0.816 | +743(-34444-35931) | 0.967 | +0.30(-0.80-1.39) | 0.594 | -17.8(-93.4-57.7) | 0.644 |
| Smoking | -0.09(-1.47) | 0.900 | -4.08(-37.1-28.9) | 0.808 | **-1.68(-3.11 to -0.26)** | **0.021** | -51.9(-711-607) | 0.877 |
| Obesity | +0.30(-0.44-1.03) | 0.425 | +86(-1679-1851) | 0.924 | -0.13(-0.72-0.46) | 0.667 | +11.9(-0.90-24.65) | 0.068 |
| Physical Inactivity | **+0.70(0.16-1.25)** | **0.012** | +10.5(-2.3-23.3) | 0.106 | +0.12(-0.44-0.68) | 0.685 | +13.46(-0.38-27.31) | 0.685 |
| Drinking | **-1.60(-2.76 to -0.45)** | **0.007** | +4.4(-2.2-10.9) | 0.189 | +0.42(-0.68-1.53) | 0.452 | -51.8(-482-379) | 0.814 |
| Uninsured(all) | **-2.85(-3.78 to -1.92)** | **<0.0001** | +0.87(-5.02-6.76) | 0.772 | **-3.73(-4.72 to -2.74)** | **<0.0001** | **+5.16(1.52-8.80)** | **0.005** |
| Some college | **+0.36(0.09-0.62)** | **0.009** | -26(-261-208) | 0.826 | **+1.08(0.81-1.35)** | **<0.0001** | **+3.83(1.17-6.48)** | **0.005** |
| Air pollution index | **+0.09(0.06-0.11)** | **<0.0001** | +0.27(-0.16-0.70) | 0.212 | **+0.12(0.10-0.13)** | **<0.0001** | **+0.06(0.02-0.11)** | **0.010** |
| HH with high cost | **+4.4(3.5-5.3)** | **<0.0001** | -44(-207-124) | 0.598 | **+5.49(4.56-6.41)** | **<0.0001** | +3.39(-2.87-9.66) | 0.288 |
| Food insecurity | **+2.72(1.78-3.67)** | **<0.0001** | +159(-783-1101) | 0.741 | +2.7(-49.4-54.8) | 0.920 | +1338(-14340-14608) | 0.986 |
| Insufficient Sleep | +0.79(-0.20-1.78) | 0.120 | -10.8(-83.8-62.2) | 0.771 | **+0.97(0.01-1.93)** | **0.047** | -6.32(-173-160) | 0.941 |
| Population age<18 | -0.78(-1.88-0.32) | 0.162 | **+25.2(3.0-46.3)** | **0.020** | -0.67(-1.90-0.56) | 0.285 | +25.5(-1.10-52.1) | 0.060 |
| Population age>65 | **+1.09(0.42-1.75)** | **0.001** | **+7.38(5.90-8.85)** | **<0.0001** | +0.42(-0.28-1.13) | 0.240 | -2.09(-14.4-10.2) | 0.739 |
| Non-Hispanic Black | **+0.47(0.14-0.79)** | **0.005** | **-7.0(-12.2 to -1.8)** | **0.008** | **+1.09(0.73-1.45)** | **<0.0001** | -5.06(-10.2-0.06) | 0.053 |
| Native Americans | **-1.1(-1.6 to -0.6)** | **<0.0001** | **-19.7(-38.5 to -1.0)** | **0.039** | **-1.68(-2.26 to -1.09)** | **<0.0001** | -49.8(-162.3-62.7) | 0.386 |
| Asians | -0.41(-2.02-1.20) | 0.614 | +4.21(-29.9-38.3) | 0.809 | **+2.47(1.02-3.92)** | **0.001** | +16.8(-12.2-45.7) | 0.256 |
| Pacific Islanders | +1.68(-2.23-5.59) | 0.401 | -192(-702-318) | 0.462 | **+10.1(6.3-13.9)** | **<0.0001** | -836(-1825-154) | 0.098 |
| Hispanics | **+0.58(0.25-0.91)** | **0.001** | **-10.7(-17.2 to -4.3)** | **0.001** | +0.57(-41.7-42.9) | 0.979 | -14.9(-2689-2660) | 0.991 |
| Non-Hispanic White | -0.11(-0.33-0.12) | 0.380 | **+2.11(1.78-2.43)** | **<0.0001** | -0.23(-0.48-0.01) | 0.065 | **+0.92(0.11-1.73)** | **0.027** |
| Female | **+7.22(5.83-8.61)** | **<0.0001** | +1.7(-18.4 -21.8) | 0.867 | **+8.46(7.07-9.86)** | **<0.0001** | **-13.2(-23.2 to -3.2)** | **0.010** |
| CVD hospitalizations | +0.002(-0.0005-0.005) | 0.100 | +0.008(-0.009-0.03) | 0.357 | +0.0008(-0.002-0.004) | 0.612 | **+0.01(0.004-0.02)** | **0.006** |
| CVD death | **+0.003(0.002-0.004)** | **<0.0001** | **+0.01(0.004-0.02)** | **0.001** | **+0.002(0.001-0.003)** | **<0.0001** | **+0.008(0.003-0.01)** | **0.003** |
| HF hospitalizations | +0.002(-0.004-0.008) | 0.563 | +0.04(-0.0007-0.08) | 0.054 | +0.001(-0.006-0.008) | 0.751 | **+0.05(0.01-0.08)** | **0.006** |
| HF death | **+0.003(0.002-0.004)** | **<0.0001** | +0.04(-0.01-0.09) | 0.127 | +0.001(-0.00006-0.003) | 0.060 | +0.04(-0.008-0.09) | 0.103 |
| CHD hospitalizations | **+0.02(0.009-0.03)** | **<0.0001** | -0.06(-0.36-0.25) | 0.717 | **+0.01(0.00001-0.02)** | **0.050** | -0.34(-3.17-2.48) | 0.812 |
| CHD death | **+0.003(0.002-0.004)** | **<0.0001** | -0.07(-0.55-0.41) | 0.779 | **+0.002(0.0008-0.003)** | **<0.0001** | +0.03(-0.004-0.07) | 0.082 |
| Diabetes | +0.0006(-0.009-0.01) | 0.893 | +0.05(-0.04-0.14) | 0.258 | -0.004(-0.01-0.006) | 0.393 | **+0.08(0.04-0.11)** | **<0.0001** |

### Covid-19 confirmed case and death rate

Covid-19 confirmed case and death rate (Figure 2) had similar geographic distributions. As of June 11^th^, 2020, in an average county, there were medians of 168.3 (IQR 67.6 – 410.9) confirmed cases and 2.95 (IQR 0 – 13.7) deaths per 100,000 population.

**Figure 2.**
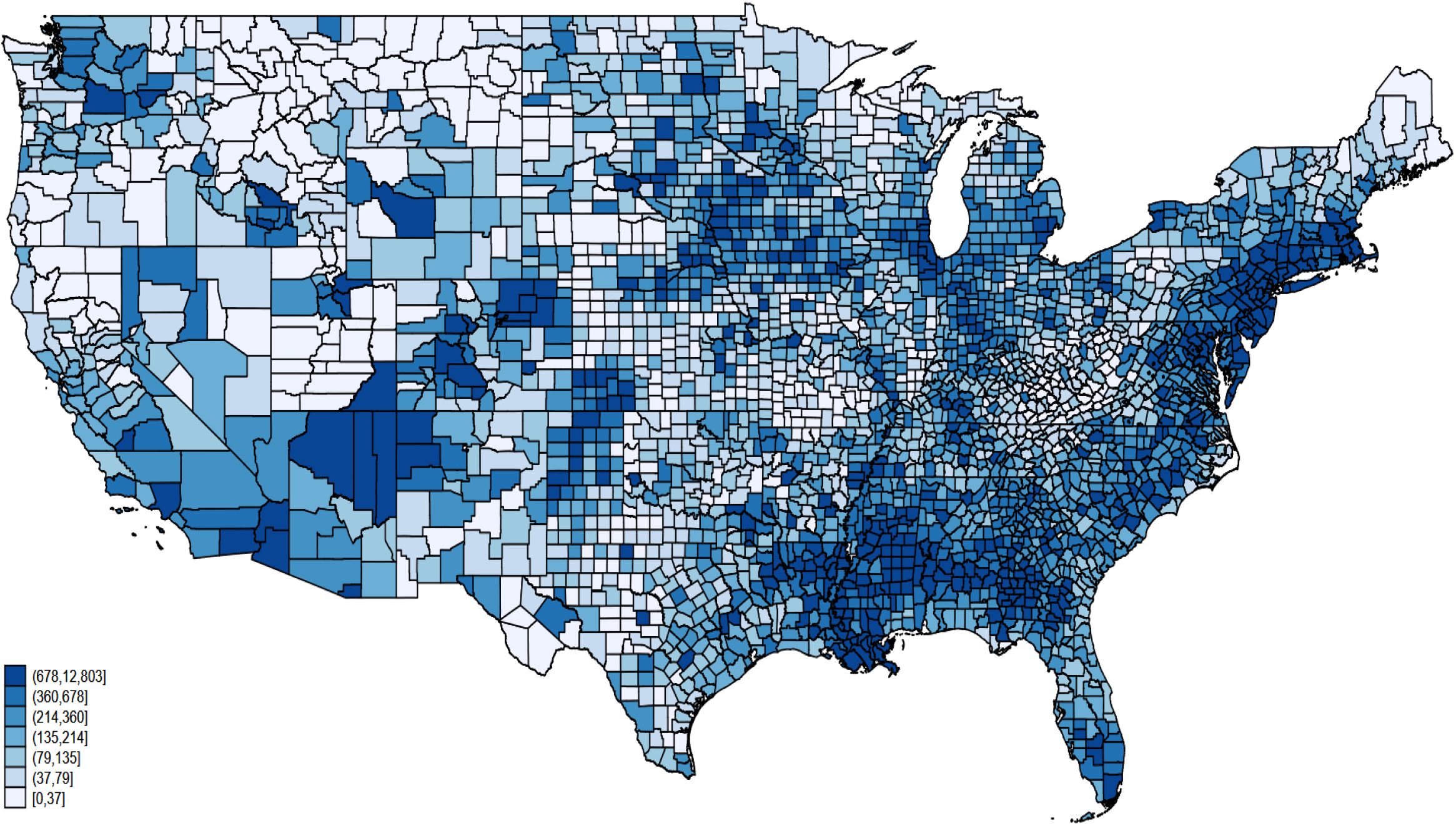
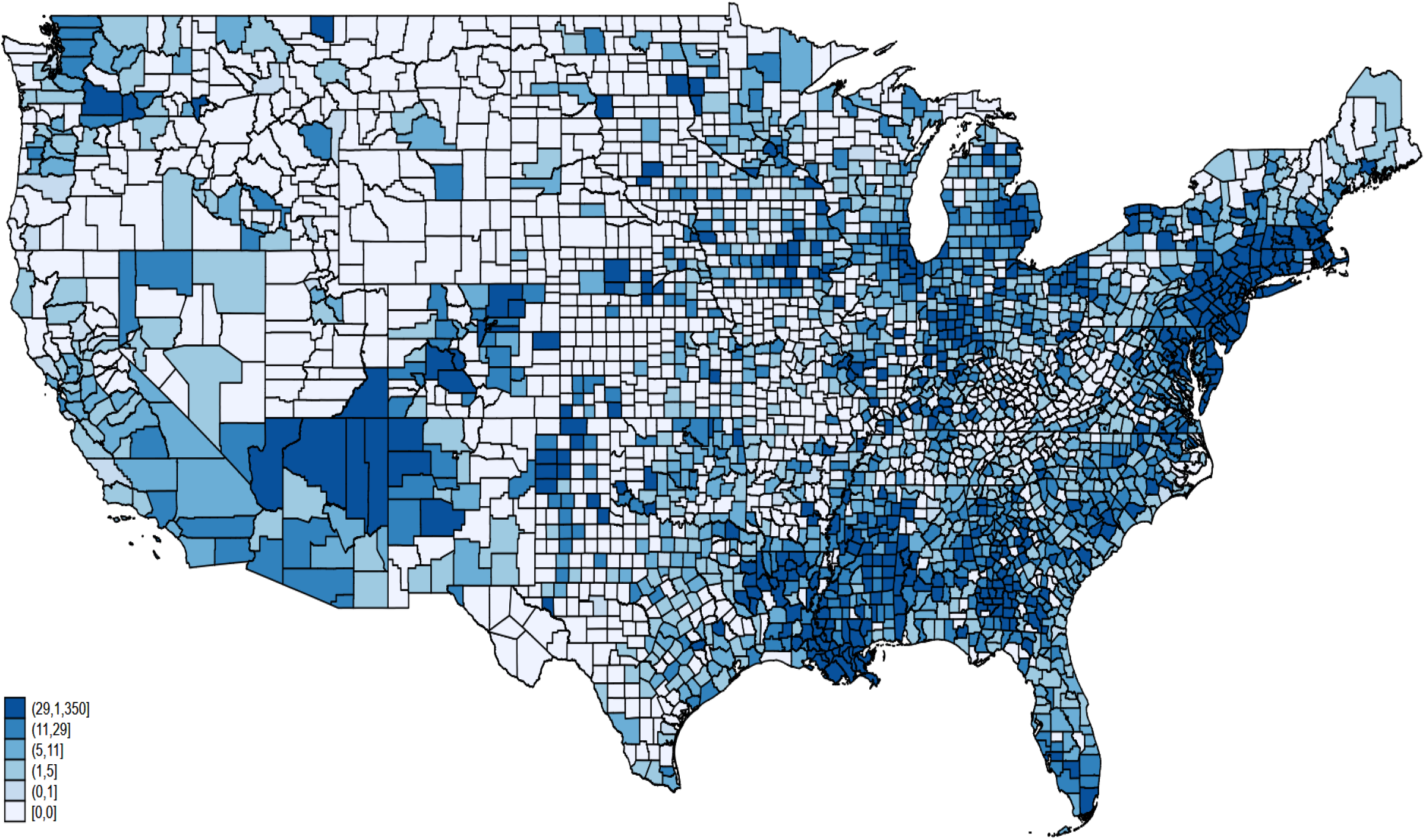
Confirmed Covid-19 cases (**A**) and deaths (**B**) in the United States adjusted for a county population size. Data of June 11^th^, 2020.

In unadjusted spatial autoregression analysis (Table 4), higher CVD and HF hospitalization rates among Medicare beneficiaries, higher percentage of Blacks, Hispanics, Asians, and Pacific Islanders, greater proportions of children (age < 18y), higher percentage of residents admitting excessive drinking, higher percentage of households with high housing costs, and worse air quality were associated with a higher rate of confirmed Covid-19 cases. In contrast, a larger proportion of county residents admitting physical inactivity, with some college education, greater percentage of adults above 65 years of age and non-Hispanic Whites, and higher CVD, HF, and total mortality across all ages were associated with a lower rate of confirmed Covid-19 cases (Table 4). The same factors that affected Covid-19 confirmed case rate also affected Covid-19 death rate, but the strength of the impact on deaths was lesser. As expected, we observed significant indirect (spillover) effects of socioeconomic factors coming from neighboring counties on confirmed Covid-19 case and death rate to a given county.

**Table 4.** Unadjusted impact on Covid-19 confirmed case and death rate

| Impact factor | Covid-19 confirmed case rate |  |  |  | Covid-19 death rate |  |  |  |
| --- | --- | --- | --- | --- | --- | --- | --- | --- |
|  | Direct (county-own) effect |  | Indirect (spillover) effect |  | Direct (county-own) effect |  | Indirect (spillover) effect |  |
| Per 1% rate increase | Marginal effect (95%CI) | P-value | Marginal effect (95%CI) | P-value | Marginal effect (95%CI) | P-value | Marginal effect (95%CI) | P-value |
| All CVD drugs use | **+0.13(0.08-0.16)** | **<0.0001** | **+0.68(0.50-0.85)** | **<0.0001** | **+0.07(0.05-0.09)** | **<0.0001** | **+0.32(0.24-0.40)** | **<0.0001** |
| Births in 2019 | **+1.22(0.90-1.54)** | **<0.0001** | **-5.60(-10.16 to -1.03)** | **<0.0001** | **+0.76(0.52-0.99)** | **<0.0001** | **-1.26(-2.32 to -0.21)** | **0.019** |
| Deaths in 2019 | **-0.48(-0.75 to -0.22)** | **<0.0001** | +1.16(-9.64-11.95) | 0.833 | **-0.23(-0.34 to -0.12)** | **<0.0001** | -0.02(-0.25-0.21) | 0.891 |
| GQ population 2019 | +0.002(-0.058-0.063) | 0.945 | -6.67(-20.8 to 7.4) | 0.353 | +0.001(-0.04-0.04) | 0.953 | **-0.53(-0.64 to -0.42)** | **<0.0001** |
| Poverty | **+0.02(0.007-0.030)** | **0.001** | **+0.50(0.29-0.70)** | **<0.0001** | +0.006(-0.004-0.02) | 0.221 | **+0.10(0.08-0.13)** | **<0.0001** |
| Median HH income | **+0.01(0.004-0.01)** | **<0.0001** | -0.17(-0.90-0.57) | 0.658 | **+0.006(0.003-0.008)** | **<0.0001** | **+0.01(0.006-0.02)** | **<0.0001** |
| Poor/Fair Health | **+5.16(3.30-7.02)** | **<0.0001** | **+36.1(18.1-54.1)** | **<0.0001** | **+2.00(0.64-3.36)** | **0.004** | **+6.89(4.74-9.04)** | **<0.0001** |
| Smoking | **+3.45(1.26-5.63)** | **0.002** | +21.7(-1.41-44.8) | 0.066 | -1.15(-2.69 to 0.40) | 0.146 | +8.77(-0.39-17.94) | 0.061 |
| Obesity | **+1.42(0.31-2.54)** | **0.012** | **+33.9(3.87-63.9)** | **0.027** | -0.82(-1.78 to 0.15) | 0.099 | **+7.11(5.13-9.09)** | **<0.0001** |
| Physical Inactivity | **-2.38(-3.46 to -1.30)** | **<0.0001** | +63.6(-38.3-165.6) | 0.221 | **-2.19(-3.17 to -1.21)** | **<0.0001** | **+10.38(7.84-12.92)** | **<0.0001** |
| Drinking | **+6.4(3.7-9.0)** | **<0.0001** | -120.1(-613.7-373.4) | 0.633 | **+2.35(0.33-4.36)** | **0.022** | **+7.67(2.42-12.93)** | **0.004** |
| Uninsured(all) | -0.20(-2.22 to 1.81) | 0.843 | -47.5(-123.3-28.2) | 0.219 | -1.15(-2.52 to 0.23) | 0.102 | **+25.9(14.3-37.6)** | **<0.0001** |
| Some college | **-0.60(-1.16 to -0.03)** | **0.037** | -3.2(-12.5-6.2) | 0.509 | **+0.77(0.33-1.21)** | **0.001** | **+2.25(1.20-3.29)** | **<0.0001** |
| Air pollution index | **+0.37(0.32-0.46)** | **<0.0001** | +0.19(-0.20-0.58) | 0.349 | **+0.27(0.23-0.32)** | **<0.0001** | -0.13(-0.33 to 0.07) | 0.195 |
| HH with high cost | **+7.3(5.4-9.2)** | **<0.0001** | **+18.4(9.9-26.8)** | **<0.0001** | **+7.78(6.30-9.26)** | **<0.0001** | +1.15(-7.37-9.66) | 0.792 |
| Food insecurity | +0.36(-1.55-2.28) | 0.710 | -145(-970.2-679.5) | 0.730 | **+2.31(0.70-3.92)** | **0.005** | **+9.86(6.49-13.22)** | **<0.0001** |
| Insufficient Sleep | **+8.8(6.5-11.0)** | **<0.0001** | +5.2(-6.4-16.9) | 0.380 | **+4.34(2.50-6.19)** | **<0.0001** | -2.79(-9.85-4.27) | 0.439 |
| Population age<18 | **+7.5(5.5-9.6)** | **<0.0001** | +76.9(-76.8-230.5) | 0.327 | **+4.36(2.93-5.79)** | **<0.0001** | +1.56(-1.26-4.38) | 0.277 |
| Population age>65 | **-9.1(-10.5 to -7.7)** | **<0.0001** | -60.7(-212.9-91.4) | 0.434 | **-5.01(-6.04 to -3.99)** | **<0.0001** | **+20.59(14.82-26.37)** | **<0.0001** |
| Non-Hispanic Black | **+2.8(2.4-3.2)** | **<0.0001** | +2.2(-1.1-5.6) | 0.190 | **+3.01(2.43-3.60)** | **<0.0001** | **+7.86(6.15-9.58)** | **<0.0001** |
| Native Americans | +0.96(-0.06-1.98) | 0.064 | **-39.5(-66.3 to -12.7)** | **0.004** | +0.30(-0.42-1.01) | 0.415 | -436.4(-1625.8-753.0) | 0.472 |
| Asians | **+5.1(1.6-8.6)** | **0.005** | **+118.6(87.7-149.5)** | **<0.0001** | **+6.48(3.99-8.96)** | **<0.0001** | **+146.2(112.8-179.6)** | **<0.0001** |
| Pacific Islanders | **+30.0(10.4-43.5)** | **0.001** | **+1245(39-2450)** | **0.043** | **+16.4(3.4-29.5)** | **0.014** | **+2751.7(351.9-5151.4)** | **0.025** |
| Hispanics | **+4.4(3.6-5.1)** | **<0.0001** | **-20.2(-34.5 to -5.9)** | **0.006** | **+2.22(1.66-2.78)** | **<0.0001** | **+27.3(8.3-46.2)** | **0.005** |
| Non-Hispanic White | **-3.1(-3.5 to -2.6)** | **<0.0001** | -0.2(-4.1 -3.6) | 0.911 | **-2.05(-2.40 to -1.70)** | **<0.0001** | **+9.09(4.47-13.72)** | **<0.0001** |
| Female | +0.3(-2.5-3.1) | 0.839 | +29.8(-30.2-89.8) | 0.330 | **+5.64(3.44-7.84)** | **<0.0001** | **-6.00(-10.77 to -1.23)** | **0.014** |
| CVD hospitalizations | **+0.02(0.01-0.02)** | **<0.0001** | -0.75(-19.5-18.0) | 0.937 | **+0.01(0.006-0.014)** | **<0.0001** | **+0.02(0.008-0.03)** | **<0.0001** |
| CVD death | -0.001(-0.002 to 0.0004) | 0.180 | +0.06(-0.03-0.16) | 0.165 | -0.001(-0.002 to 0.0001) | 0.092 | **+0.009(0.007-0.01)** | **<0.0001** |
| HF hospitalizations | **+0.05(0.04-0.06)** | **<0.0001** | -0.01(-0.16-0.13) | 0.862 | **+0.04(0.02-0.05)** | **<0.0001** | +0.06(-.10-0.22) | 0.471 |
| HF death | **-0.003(-0.005 to -0.006)** | **0.015** | -0.08(-0.20-0.04) | 0.174 | **-0.003(-0.005 to -0.001)** | **0.011** | **+0.03(0.02-0.03)** | **<0.0001** |
| CHD hospitalizations | +0.01(-0.01-0.03) | 0.216 | +0.21(-1.33-1.75) | 0.788 | -0.03(-0.05-0.01) | 0.178 | +0.01(-1.82-1.85) | 0.988 |
| CHD death | **-0.002(-0.004 to -0.0002)** | **0.028** | +0.30(-0.30-0.91) | 0.324 | **-0.003(-0.005 to -0.002)** | **<0.0001** | **+0.03(0.02-0.03)** | **<0.0001** |
| Diabetes | **+0.02(0.007-0.04)** | **0.006** | +0.46(0.07-0.83) | 0.016 | +0.006(-0.01-0.02) | 0.406 | +0.10(-0.20-0.40) | 0.524 |

HH=household

### Association of ACEI and ARB use rate with confirmed Covid-19 case and death rate

In unadjusted analysis, as expected, both ACEI and ARB use rate had a direct and indirect impact on confirmed Covid-19 case and death rate (Table 5). In bivariate analysis, only ARB but not ACEI use was associated with higher rate of confirmed cases. The association of ACEI use with Covid-19 was fully explained by confounders (Table 5). In adjusted analysis, the ACEI use rate had no direct or indirect impact on Covid-19 confirmed case rate, whereas the ARB use was associated with higher rate of confirmed Covid-19 cases. On average, an increase in ARB use rate in a given county by 1% was associated with an increase in Covid-19 confirmed case rate by 0.12%.

**Table 5.** Unadjusted and adjusted impact of the ACEIs and ARBs use rate on Covid-19 confirmed case and death rate

|  |  | Confirmed case rate |  |  |  | Death rate |  |  |  |
| --- | --- | --- | --- | --- | --- | --- | --- | --- | --- |
| Impact factor |  | Direct (county-own) effect |  | Indirect (spillover) effect |  | Direct (county-own) effect |  | Indirect (spillover) effect |  |
| Model |  | Marginal effect (95%CI) | P-value | Marginal effect (95%CI) | P-value | Marginal effect (95%CI) | P-value | Marginal effect (95%CI) | P-value |
| U | ACEI | **+0.14(0.09-0.18)** | **<0.0001** | **+1.02(0.66-1.38)** | **<0.0001** | **+0.07(0.04-0.09)** | **<0.0001** | **+0.60(0.16-1.04)** | **0.007** |
| U | ARB | **+0.14(0.10-0.18)** | **<0.0001** | **+1.18(0.82-1.54)** | **<0.0001** | **+0.09(0.07-0.11)** | **<0.0001** | **+0.91(0.71-1.11)** | **<0.0001** |
| U | Pure ARB | **+0.14(0.10-0.18)** | **<0.0001** | **+1.18(0.82-1.54)** | **<0.0001** | **+0.09(0.07-0.11)** | **<0.0001** | **+0.90(0.70-1.10)** | **<0.0001** |
| A | ACEI | +0.11(-0.31-0.53) | 0.600 | -0.53(-3.89-2.84) | 0.760 | **+0.06(0.006-0.11)** | **0.027** | +0.03(-0.22-0.28) | 0.824 |
| A | ARB | **+0.12(0.05-0.19)** | **0.002** | -0.33(-2.11-1.44) | 0.714 | **+0.06(0.04-0.09)** | **<0.0001** | +0.11(-0.20-0.41) | 0.499 |
| A | Pure ARB | **+0.12(0.01-0.22)** | **0.028** | -0.27(-3.40-2.87) | 0.868 | **+0.06(0.04-0.09)** | **<0.0001** | +0.11(-0.19-0.42) | 0.465 |
| B | ACEI | +0.005(-0.13-0.14) | 0.936 | -6.77(-17.65-4.10) | 0.222 | **-0.12(-0.19 to -0.06)** | **<0.0001** | **+2.58(0.12-5.04)** | **0.040** |
| B | ARB | **+0.14(0.01-0.26)** | **0.030** | +8.53(-2.83-19.89) | 0.141 | **+0.21(0.15-0.27)** | **<0.0001** | -2.33(-5.27-0.61) | 0.120 |

U=unadjusted. A=adjusted for the percentage of Black non-Hispanic county residents, percentage of a county residents younger
than 18 years of age, percentage of residents with at least some college degree, median household income as a percent of the state total, air quality index, CDC-reported CVD hospitalization rate in Medicare beneficiaries, and CVD death rate in a total county population. B=bivariate model including two predictors (ACEI and ARB use rate).

In unadjusted analysis, both ACEI and ARB use rate was associated with Covid-19 death rate. In the bivariate model, ACEI and ARB use rates demonstrated the opposite direct (county- own) impact on the death rate. Higher ACEI use rate was associated with a lower death rate, whereas higher ARB use rate was associated with higher death rate (Table 5). After adjustment for demographic and socioeconomic characteristics, air quality index, and CVD prevalence and mortality, the rates of ACEI and ARB use remained associated with Covid-19 death rate. On average, an increase in ARB use rate in a given county by 1% was associated with a 0.06% increase in Covid-19 deaths in that county (direct, county-own effect).

Of note, adjustment fully explained the indirect (spillover) impact of ACEI and ARB use rate coming from all neighboring counties on Covid-19 confirmed case and death rate to a given county, while all models confirmed strong spatial dependence (Moran test of spatial terms for all models *P*<0.00001). The marginal analysis showed no significant differences in outcomes if ACEI and ARB use rate would change in the same direction (either increase or decrease) in all counties (Supplemental Figure 1).

### Sensitivity analysis

In unadjusted analysis, the use of cardiovascular medications reflects CVD prevalence. As expected, the use of all types of cardiovascular medications was associated with Covid-19 confirmed case rate (Supplemental Table 1). Overall, the association with the Covid-19 death rate was similar but had a smaller effect size. The effect size for ACEI and ARB use rate was comparable to that of other cardiovascular medications.

In adjusted analysis (Supplemental Table 2), the rate of use of the vast majority of cardiovascular medications had no association with Covid-19 confirmed case rate, with very few exceptions. This finding suggests an absence of significant reverse causality bias and allows meaningful interpretation of the main study results for the primary outcome.

Similarly to the main results, after adjustment, most of the cardiovascular mediations remained significantly associated with Covid-19 death rate (Supplemental Table 2). Therefore, reverse causality bias was likely present in our analyses with the Covid-19 death rate outcome, which limited meaningful interpretation of the main results for the secondary outcome.

Results of the analyses with pure ARB class of medications did not differ from the main study results (Table 5).

## Discussion

Our study confirmed that the rate of ACEI use does not impact Covid-19 confirmed case rate. However, we observed that ARB use associated with slightly increased Covid-19 confirmed case rates. After adjustment for demographic and socioeconomic confounders and CVD prevalence and severity, an increase in ARB use by 1% was associated with a 0.12 % increase in Covid-19 confirmed cases. These findings suggest that long-term use of ARB, due to known ACE2 upregulation, may facilitate SARS-CoV-2 entry into target cells and increase infectivity. ACEI and ARB are frequently prescribed interchangeably for the same clinical indications. Cluster-randomized controlled trial is warranted to answer the question of whether the replacement of ARB by ACEI may reduce the Covid-19 confirmed case rate. Our results highlight the safety and indicate possible benefits of ACEI use for patients with clinical indications for ACEI in the Covid-19 era, consistently with several other studies of ACEI in Covid-19.^28,29^

Importantly, in this observational geospatial study, residual confounding and reverse causality bias cannot be completely ruled out. The use of ARB may indicate a subgroup of CVD patients who are especially vulnerable to the virus. In such a case, the use of ARBs is not a cause, but a marker of risk. Reverse causality bias likely explains an association of ACEI and ARB use with the Covid-19 death rate, which was similar to the vast majority of other cardiovascular medications.

### SARS-CoV-2 virus may preferentially infect individuals taking ARB, but not ACEI

There is strong evidence that the entry of the SARS-CoV-2 virus into the host cell depends on the SARS-CoV receptor ACE2. ACE2 is a type I integral membrane glycoprotein expressed mainly in the respiratory tract, heart, kidneys, and gastrointestinal tract. ACE2 tissue expression facilitates the virus entry in target cells.^5^

Clinical indications for ACEI and ARB are similar, and in many clinical studies, these two classes of drugs considered together under the common name “RAAS inhibitors.” However, the effects of ACEI and ARB on ACE2 levels and activity are different.^3^ Our findings support the hypothesis that long-term use of ARB, but not ACEI may facilitate SARS-CoV-2 entry and increase infectivity.

Several unmeasured confounders could be responsible for our findings. First, ARB are indicated for patients who are intolerant to ACEI, usually because of bradykinin-mediated cough. Second, both ACEI and ARB can be used together in patients with advanced HF, and thus indicate a high-risk patient population. For those patient categories, switching from ARB to ACEI is not an option. Nevertheless, regardless of whether ARB indeed increases infectivity or simply indicates a high-risk patient population, it would be wise to reinforce effective Covid-19 prevention strategies, to minimize the risks of exposure to the virus.

Our findings support the notion of ACEI and ARB playing a double-edged sword role. The strength of associations of ACEI and ARB use with the Covid-19 death rate was twice less than with confirmed case rate, suggesting no adverse effect on Covid-19 disease severity.

### Limitations

Although the Medicare Part D Prescriber Public Use File has a wealth of information, the dataset has several limitations. The data may not be representative of a physician’s entire practice or all of Medicare as it only includes information on beneficiaries enrolled in the Medicare Part D prescription drug program (approximately two-thirds of all Medicare beneficiaries). Besides, available data were for the year 2017 and did not reflect the most recent use of medications in 2020. Nevertheless, we measured exposure before the outcome, which is essential for the interpretation of the study findings. Furthermore, we did not adjust for adherence to medications. Nevertheless, a recent geospatial study of ACEI/ARB adherence^30^ showed a relatively consistent geographic distribution of ACEI/ARB adherence across the US. An observational cross-sectional geospatial study is susceptible to reverse causality bias. To address this limitation, we performed a rigorous analysis of all other classes of cardiovascular medications. The results of sensitivity analyses helped to identify the reverse causality bias in analyses with the secondary outcome. Finally, unobserved confounding was likely present in this observational study. The most apparent missing data included the rate of Covid-19 testing. Therefore, observed effect sizes have to be interpreted with caution. However, unobserved confounding would unlikely to affect a relative comparison of ACEI and ARB.

## Data Availability

We provide the study dataset and STATA (StataCorp, College Station, Texas) code at https://github.com/Tereshchenkolab/geospatial.

## Acknowledgments

Authors acknowledge numerous scientists and institutions that provided open data.

---

**Supplemental Table 1.**
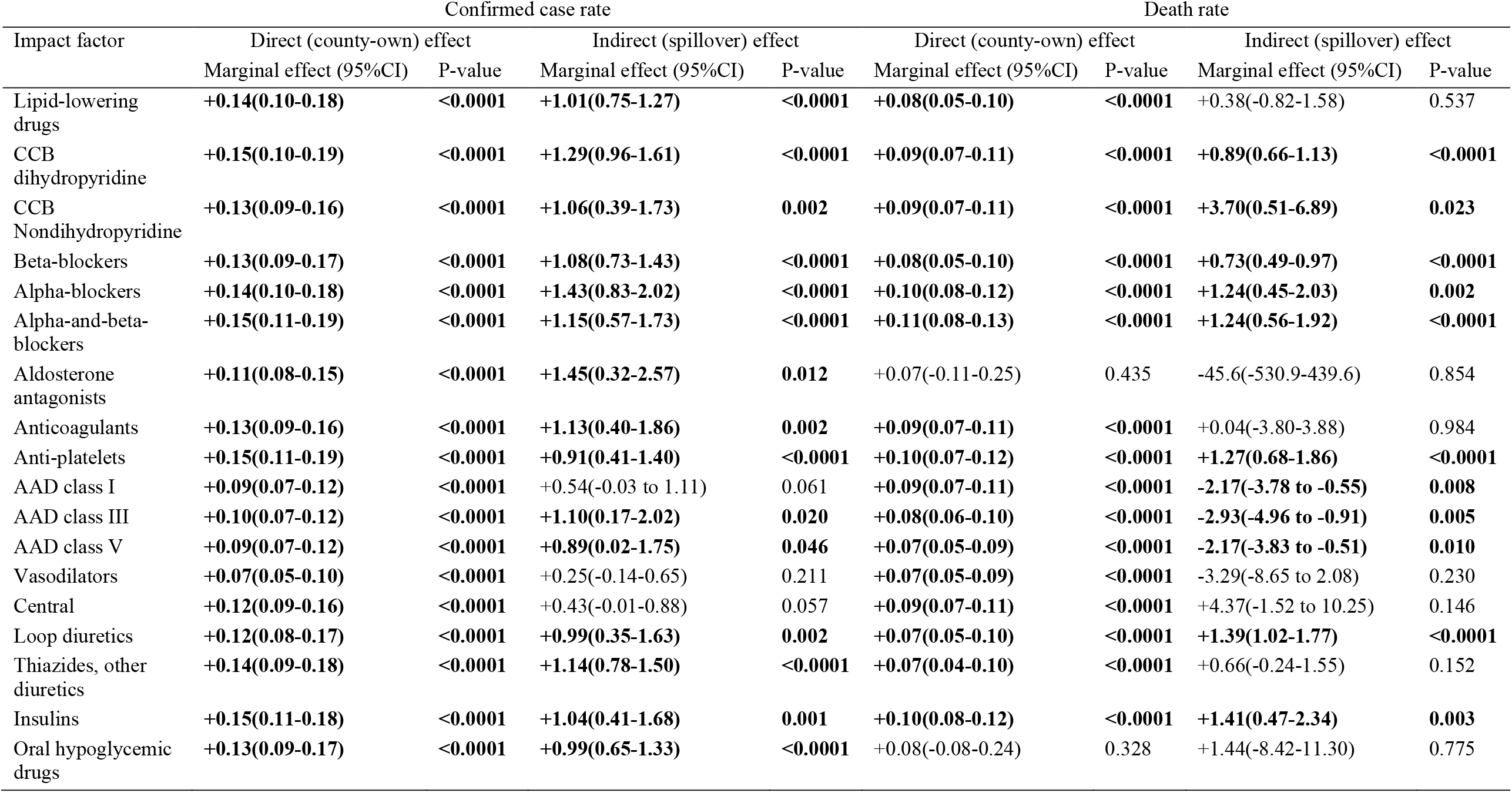
Unadjusted impact of the use of cardiovascular medications on Covid-19 confirmed case and death rate

| Impact factor | Confirmed case rate |  |  |  | Death rate |  |  |  |
| --- | --- | --- | --- | --- | --- | --- | --- | --- |
|  | Direct (county-own) effect |  | Indirect (spillover) effect |  | Direct (county-own) effect |  | Indirect (spillover) effect |  |
|  | Marginal effect (95%CI) | P-value | Marginal effect (95%CI) | P-value | Marginal effect (95%CI) | P-value | Marginal effect (95%CI) | P-value |
| Lipid-lowering drugs | **+0.14(0.10-0.18)** | **<0.0001** | **+1.01(0.75-1.27)** | **<0.0001** | **+0.08(0.05-0.10)** | **<0.0001** | +0.38(-0.82-1.58) | 0.537 |
| CCB dihydropyridine | **+0.15(0.10-0.19)** | **<0.0001** | **+1.29(0.96-1.61)** | **<0.0001** | **+0.09(0.07-0.11)** | **<0.0001** | **+0.89(0.66-1.13)** | **<0.0001** |
| CCB Nondihydropyridine | **+0.13(0.09-0.16)** | **<0.0001** | **+1.06(0.39-1.73)** | **0.002** | **+0.09(0.07-0.11)** | **<0.0001** | **+3.70(0.51-6.89)** | **0.023** |
| Beta-blockers | **+0.13(0.09-0.17)** | **<0.0001** | **+1.08(0.73-1.43)** | **<0.0001** | **+0.08(0.05-0.10)** | **<0.0001** | **+0.73(0.49-0.97)** | **<0.0001** |
| Alpha-blockers | **+0.14(0.10-0.18)** | **<0.0001** | **+1.43(0.83-2.02)** | **<0.0001** | **+0.10(0.08-0.12)** | **<0.0001** | **+1.24(0.45-2.03)** | **0.002** |
| Alpha-and-beta-blockers | **+0.15(0.11-0.19)** | **<0.0001** | **+1.15(0.57-1.73)** | **<0.0001** | **+0.11(0.08-0.13)** | **<0.0001** | **+1.24(0.56-1.92)** | **<0.0001** |
| Aldosterone antagonists | **+0.11(0.08-0.15)** | **<0.0001** | **+1.45(0.32-2.57)** | **0.012** | +0.07(-0.11-0.25) | 0.435 | -45.6(-530.9-439.6) | 0.854 |
| Anticoagulants | **+0.13(0.09-0.16)** | **<0.0001** | **+1.13(0.40-1.86)** | **0.002** | **+0.09(0.07-0.11)** | **<0.0001** | +0.04(-3.80-3.88) | 0.984 |
| Anti-platelets | **+0.15(0.11-0.19)** | **<0.0001** | **+0.91(0.41-1.40)** | **<0.0001** | **+0.10(0.07-0.12)** | **<0.0001** | **+1.27(0.68-1.86)** | **<0.0001** |
| AAD class I | **+0.09(0.07-0.12)** | **<0.0001** | +0.54(-0.03 to 1.11) | 0.061 | **+0.09(0.07-0.11)** | **<0.0001** | **-2.17(-3.78 to -0.55)** | **0.008** |
| AAD class III | **+0.10(0.07-0.12)** | **<0.0001** | **+1.10(0.17-2.02)** | **0.020** | **+0.08(0.06-0.10)** | **<0.0001** | **-2.93(-4.96 to -0.91)** | **0.005** |
| AAD class V | **+0.09(0.07-0.12)** | **<0.0001** | **+0.89(0.02-1.75)** | **0.046** | **+0.07(0.05-0.09)** | **<0.0001** | **-2.17(-3.83 to -0.51)** | **0.010** |
| Vasodilators | **+0.07(0.05-0.10)** | **<0.0001** | +0.25(-0.14-0.65) | 0.211 | **+0.07(0.05-0.09)** | **<0.0001** | -3.29(-8.65 to 2.08) | 0.230 |
| Central | **+0.12(0.09-0.16)** | **<0.0001** | +0.43(-0.01-0.88) | 0.057 | **+0.09(0.07-0.11)** | **<0.0001** | +4.37(-1.52 to 10.25) | 0.146 |
| Loop diuretics | **+0.12(0.08-0.17)** | **<0.0001** | **+0.99(0.35-1.63)** | **0.002** | **+0.07(0.05-0.10)** | **<0.0001** | **+1.39(1.02-1.77)** | **<0.0001** |
| Thiazides, other diuretics | **+0.14(0.09-0.18)** | **<0.0001** | **+1.14(0.78-1.50)** | **<0.0001** | **+0.07(0.04-0.10)** | **<0.0001** | +0.66(-0.24-1.55) | 0.152 |
| Insulins | **+0.15(0.11-0.18)** | **<0.0001** | **+1.04(0.41-1.68)** | **0.001** | **+0.10(0.08-0.12)** | **<0.0001** | **+1.41(0.47-2.34)** | **0.003** |
| Oral hypoglycemic drugs | **+0.13(0.09-0.17)** | **<0.0001** | **+0.99(0.65-1.33)** | **<0.0001** | +0.08(-0.08-0.24) | 0.328 | +1.44(-8.42-11.30) | 0.775 |

**Supplemental Table 2.**
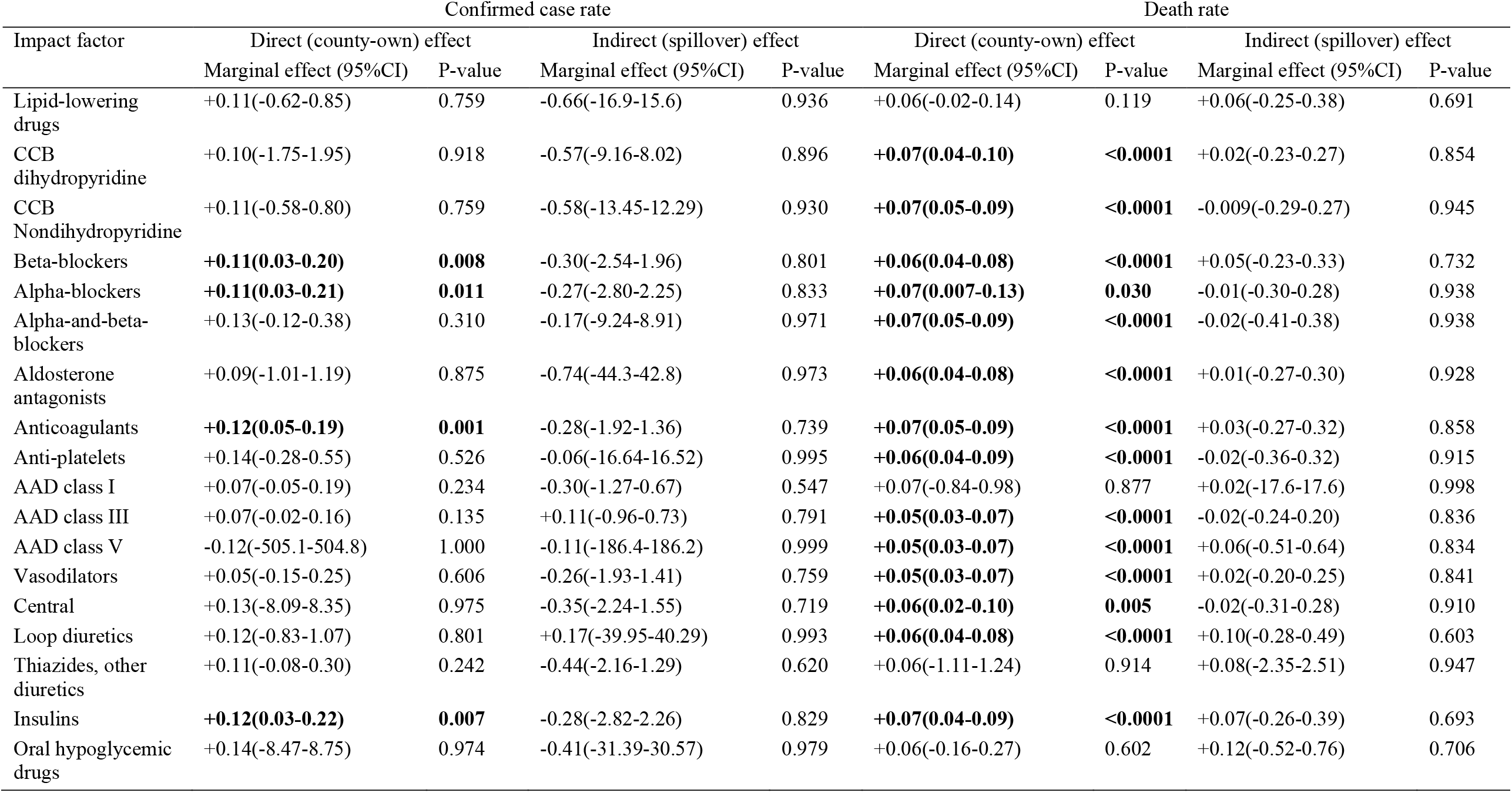
Adjusted impact of cardiovascular medications use on Covid-19 confirmed case and death rate

**Supplemental Figure 1:**
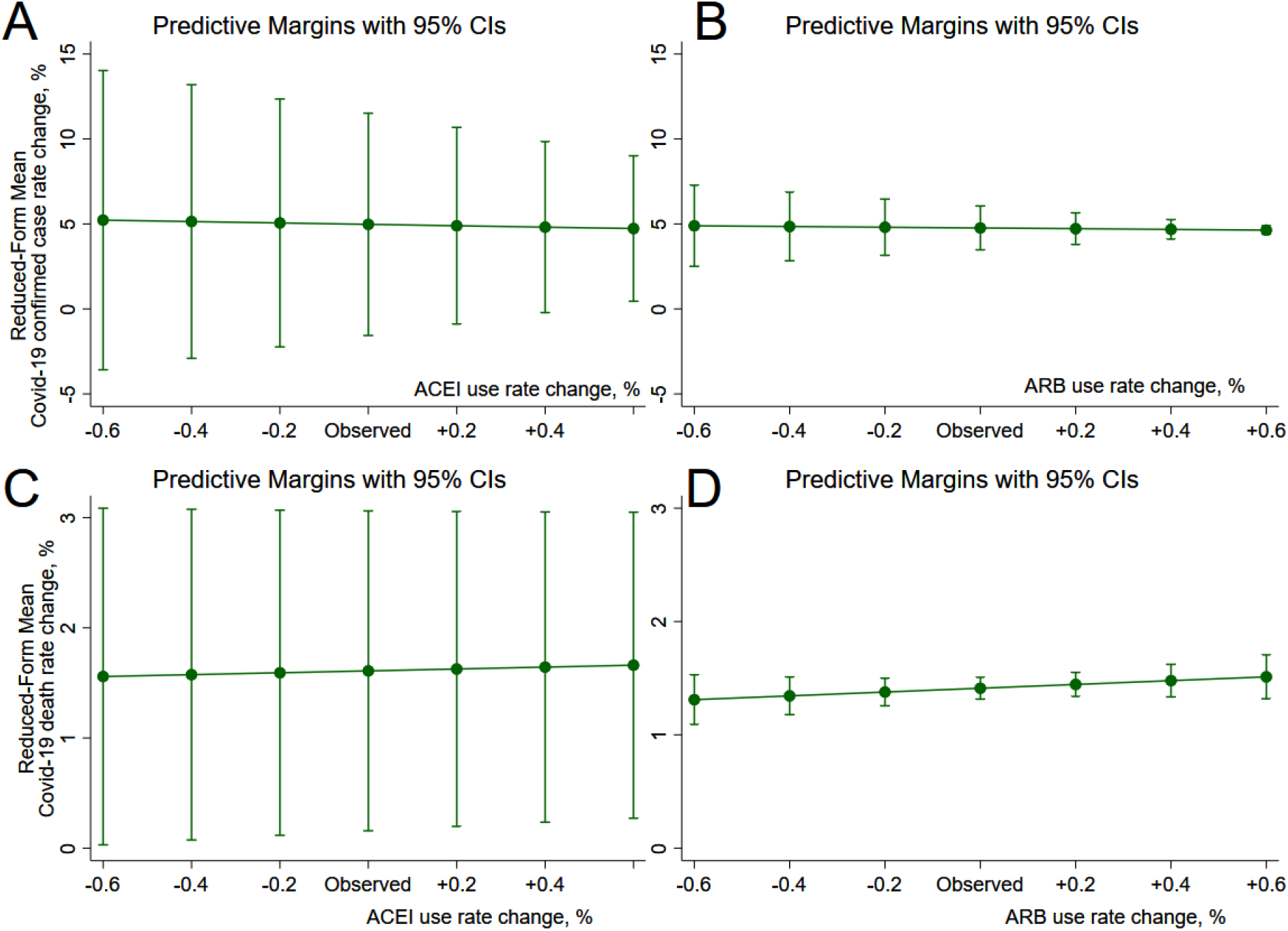
Marginsplot of the adjusted effect of ACEI (A, C) and ARB (B, D) use rate on Covid-19 confirmed cases rate (A, B) and death rate (C, D). All variables were log-transformed and reflected the relative change. Plots show how would Covid-19 confirmed case rate and death rate change if ACEI and ARB use rate in all counties either drop or increase by 0.1 – 0.6 %. All models were adjusted for the percentage of Black non-Hispanic county residents, percentage of a county residents younger than 18 years of age, percentage of residents with at least some college degree, median household income as a percent of the state total, air quality index, CVD hospitalization rate in Medicare beneficiaries, and CVD death rate in a total county population. Total impact is shown, as a sum of direct and indirect impacts.

## References

1. Zhou P, Yang XL, Wang XG, et al. A pneumonia outbreak associated with a new coronavirus of probable bat origin. Nature 2020; 579(7798): 270–3.

2. Kuba K, Imai Y, Rao S, et al. A crucial role of angiotensin converting enzyme 2 (ACE2) in SARS coronavirus-induced lung injury. Nat Med 2005; 11(8): 875–9.

3. Vaduganathan M, Vardeny O, Michel T, McMurray JJV, Pfeffer MA, Solomon SD. Renin-Angiotensin-Aldosterone System Inhibitors in Patients with Covid-19. N Engl J Med 2020; 382(17): 1653–9.

4. Zhou F, Yu T, Du R, et al. Clinical course and risk factors for mortality of adult inpatients with COVID-19 in Wuhan, China: a retrospective cohort study. Lancet 2020; 395(10229): 1054–62.

5. Hofmann H, Geier M, Marzi A, et al. Susceptibility to SARS coronavirus S protein-driven infection correlates with expression of angiotensin converting enzyme 2 and infection can be blocked by soluble receptor. Biochem Biophys Res Commun 2004; 319(4): 1216–21.

6. Ishiyama Y, Gallagher PE, Averill DB, Tallant EA, Brosnihan KB, Ferrario CM. Upregulation of angiotensin-converting enzyme 2 after myocardial infarction by blockade of angiotensin II receptors. Hypertension 2004; 43(5): 970–6.

7. Soler MJ, Ye M, Wysocki J, William J, Lloveras J, Batlle D. Localization of ACE2 in the renal vasculature: amplification by angiotensin II type 1 receptor blockade using telmisartan. Am J Physiol Renal Physiol 2009; 296(2): F398–405.

8. Sukumaran V, Veeraveedu PT, Gurusamy N, et al. Cardioprotective effects of telmisartan against heart failure in rats induced by experimental autoimmune myocarditis through the modulation of angiotensin-converting enzyme-2/angiotensin 1-7/mas receptor axis. Int J Biol Sci 2011; 7(8): 1077–92.

9. Lakshmanan AP, Thandavarayan RA, Watanabe K, et al. Modulation of AT-1R/MAPK cascade by an olmesartan treatment attenuates diabetic nephropathy in streptozotocin-induced diabetic mice. Molecular and Cellular Endocrinology 2012; 348(1): 104–11.

10. Igase M, Strawn WB, Gallagher PE, Geary RL, Ferrario CM. Angiotensin II AT1 receptors regulate ACE2 and angiotensin-(1-7) expression in the aorta of spontaneously hypertensive rats. Am J Physiol Heart Circ Physiol 2005; 289(3): H1013–9.

11. Zhong JC, Ye JY, Jin HY, et al. Telmisartan attenuates aortic hypertrophy in hypertensive rats by the modulation of ACE2 and profilin-1 expression. Regul Pept 2011; 166(1-3): 90–7.

12. Sukumaran V, Tsuchimochi H, Tatsumi E, Shirai M, Pearson JT. Azilsartan ameliorates diabetic cardiomyopathy in young db/db mice through the modulation of ACE-2/ANG 1-7/Mas receptor cascade. Biochemical pharmacology 2017; 144: 90–9.

13. Ferrario CM, Jessup J, Chappell MC, et al. Effect of angiotensin-converting enzyme inhibition and angiotensin II receptor blockers on cardiac angiotensin-converting enzyme 2. Circulation 2005; 111(20): 2605–10.

14. Burrell LM, Risvanis J, Kubota E, et al. Myocardial infarction increases ACE2 expression in rat and humans. European Heart Journal 2005; 26(4): 369–75.

15. Hamming I, Van Goor H, Turner AJ, et al. Differential regulation of renal angiotensin-converting enzyme (ACE) and ACE2 during ACE inhibition and dietary sodium restriction in healthy rats. Experimental Physiology 2008; 93(5): 631–8.

16. Rice GI, Thomas DA, Grant PJ, Turner AJ, Hooper NM. Evaluation of angiotensin-converting enzyme (ACE), its homologue ACE2 and neprilysin in angiotensin peptide metabolism. Biochem J 2004; 383(Pt 1): 45–51.

17. United States Census Bureau. Cartographic Boundary Files - Shapefile. 2018. https://www.census.gov/geographies/mapping-files/time-series/geo/carto-boundary-file.html. Accessed May 5, 2020.

18. The Medicare Provider Utilization and Payment Data: Part D Prescriber Public Use File: 2017. https://data.cms.gov/Medicare-Part-D/Medicare-Provider-Utilization-and-Payment-Data-201/77gb-8z53/data. Accessed 5/18/2020.

19. Google Geocoding API. Googles Maps Platform. https://developers.google.com/maps/documentation/geocoding/start. Accessed 5/18/2020.

20. Dong E, Du H, Gardner L. An interactive web-based dashboard to track COVID-19 in real time. Lancet Infect Dis 2020; 20(5): 533–4.

21. U.S. Census Bureau, Population Division. CO-EST2019-alldata: Annual Resident Population Estimates, Estimated Components of Resident Population Change, and Rates of the Components of Resident Population Change for States and Counties: April 1, 2010 to July 1, 2019. File: 7/1/2019 County Population Estimates. Release Date: March 2020. https://www2.census.gov/programs-surveys/popest/datasets/2010-2019/counties/totals/. Group Quarters Information. https://www.census.gov/2018censustest/gq. Accessed 5/18/2020.

22. The United States Department of Agriculture. Economic Research Service. County-Level Data Sets. https://www.ers.usda.gov/data-products/county-level-data-sets/. Accessed 5/18/2020.

23. The Robert Wood Johnson Foundation and the University of Wisconsin Population Health Institute. County Health Rankings & Roadmaps program. https://www.countyhealthrankings.org/explore-health-rankings/rankings-data-documentation. Accesssed 5/18/2020.

24. The Centers for Disease Control and Prevention. National Center for Chronic Disease Prevention and Health Promotion, Division for Heart Disease and Stroke Prevention. https://www.cdc.gov/dhdsp/maps/atlas/data-sources.html#dataControls. https://nccd.cdc.gov/DHDSPAtlas/?state=County&tt=HI&classid=1&subid=1&hifilters=%5B%5B9,1%5D,%5B2,1%5D,%5B3,1%5D,%5B4,1%5D,%5B7,1%5D%5D&ol=%5B10,14%5D. Accessed 5/18/2020.

25. Drukker DM, Egger P, Prucha IR. On Two-Step Estimation of a Spatial Autoregressive Model with Autoregressive Disturbances and Endogenous Regressors. Econometric Reviews 2013; 32(5-6): 686-733.

26. Lee L-F. Asymptotic Distributions of Quasi-Maximum Likelihood Estimators for Spatial Autoregressive Models. Econometrica 2004; 72(6): 1899–925.

27. Kelejian HH, Prucha IR. Specification and estimation of spatial autoregressive models with autoregressive and heteroskedastic disturbances. Journal of Econometrics 2010; 157(1): 53–67.

28. de Abajo FJ, Rodríguez-Martín S, Lerma V, et al. Use of renin–angiotensin–aldosterone system inhibitors and risk of COVID-19 requiring admission to hospital: a case-population study. The Lancet 2020.

29. Bean D, Kraljevic Z, Searle T, et al. ACE-inhibitors and Angiotensin-2 Receptor Blockers are not associated with severe SARS-COVID19 infection in a multi-site UK acute Hospital Trust. medRxiv 2020: 2020.04.07.20056788.

30. Han Y, Saran R, Erickson SR, Hirth RA, He K, Balkrishnan R. Environmental and individual predictors of medication adherence among elderly patients with hypertension and chronic kidney disease: A geospatial approach. Research in Social and Administrative Pharmacy 2020; 16(3): 422–30.

